# Constipation is associated with an increased risk of major adverse cardiac events in a UK population

**DOI:** 10.1101/2024.01.22.24301635

**Authors:** Tenghao Zheng, Leticia Camargo Tavares, Mauro D’Amato, Francine Z. Marques

## Abstract

**Background:** Traditional cardiovascular risk factors, including hypertension, only explain part of major adverse cardiac events (MACE). Understanding what other risk factors contribute to MACE is essential for prevention. Constipation shares common risk factors with hypertension and is associated with an increased risk of several cardiovascular diseases. We hypothesised that constipation is an under-appreciated risk factor for MACE.

**Methods:** We used the population healthcare and genomic data in the UK Biobank (UKBB) (n=408,354) to study the contribution of constipation (ICD-10 K59.0) to the risk of MACE, defined by any episode of acute coronary syndrome (ACS), ischemic stroke and heart failure (HF). Analyses were controlled for traditional cardiovascular risk factors. We also assessed genetic correlations (r_g_) between constipation and MACE.

**Results:** Constipation cases (N=23,814) exhibited significantly higher risk of MACE compared to those with normal bowel habits (OR=2.15, P<1.00×10^-300^). Constipation was also significantly associated with individual MACE subgroups, in order: HF (OR=2.72, P<1.00×10^-300^), ischemic stroke (OR=2.36, P=2.02×10^-230^), and ACS (OR=1.62, P=5.82×10^-113^). In comparison with constipation-free hypertensive patients, hypertensive patients with constipation showed significantly higher odds of MACE (OR=1.68, P=1.05×10^-136^) and a 34% increased risk of death (P=2.3×10^-50^) after adjustment for medications that affect gut motility and other traditional cardiovascular risk factors. Finally, we detected positive genetic correlations between constipation and MACE subgroups ACS (r_g_=0.27, P=2.12×10^-6^), ischemic stroke (r_g_ =0.23, P=0.011), and HF (r_g_ =0.21, P=0.0062).

**Conclusion:** We identified constipation as a potential risk factor independently associated with higher MACE prevalence. These findings warrant further studies on their causal relationship and identification of pathophysiological mechanisms.

## Introduction

Cardiovascular diseases (CVD) are the leading global cause of death, primarily attributed to major adverse cardiac events (MACE) such as ischemic stroke, acute coronary syndrome (ACS), and heart failure (HF).^1,2^ High blood pressure (BP), known as hypertension, is a key modifiable risk factor for MACE.^2-4^ However, hypertension and other traditional metabolic (obesity, diabetes, high cholesterol levels), demographical (older age, male sex), and behavioural (smoking) factors identified so far can only explain part of the risk.^5,6^ Hence, identifying of other non-traditional risk factors contributing to MACE is crucial to improve prevention, discover new therapeutic interventions, and implement more effective management strategies based on individual’s risk assessment, in line with precision medicine principles.

Recently, connections between the gut, heart and brain have been recognised,^7,8^ with accumulating evidence indicating that reduced bowel movement (constipation) is associated with an increased risk of hypertension and CVD.^9-13^ Constipation is one of the most common gastrointestinal (GI) disorders, estimated to chronically impair 14% of the general population (varying with the adopted definition), with prevalence increasing with age and affecting mostly women.^14^ Besides ageing, gut dysmotility that is observed in constipation shares common risk factors with hypertension and CVDs. Examples include autonomic nervous system dysregulation, gut dysbiosis, low dietary fibre and fluid intake, and lack of physical activity.^15-25^ A study of >540,000 hospitalised and older than 60 years Australian patients reported that, in comparison to age-matched non-constipated and non-hypertensive individuals, constipated individuals had higher risk of hypertension and cardiovascular events.^10^ It is unclear if these results apply to healthy individuals outside a hospital setting. Similarly, a Danish population-based study of >900,000 individuals found constipated individuals had an increased risk for several CVDs, particularly splanchnic venous thromboembolism.^9^

These studies, however, have some limitations. For example, antihypertensive medications, such as calcium channel blockers (CCBs), can cause constipation as a side effect.^26,27^ CCB agents (e.g., verapamil) can alter GI function and motility by decreasing smooth muscle contraction and inhibiting cellular secretion.^27^ However, it is unclear if constipation was independently associated with hypertension and CVD, or was a result of medication. Shared pathophysiological mechanisms likely play a role in constipation, hypertension, and CVD. Evidence linking constipation and CVD also comes from analyses at the genetic level. Loss-of-function genetic mutations in the ion channel gene *SCN5A* (coding for the NaV1.5 sodium channel), which plays pacemaker roles in both the heart and GI tract, have been associated with cardiac arrythmias and the irritable bowel syndrome in the constipation subtype (IBS-C).^28-30^

Hence, we hypothesised that constipation might be an under-appreciated risk factor for high BP and MACE. Herein, we investigated the contribution of constipation to hypertension and MACE, and the genetic predisposition behind it in a large population-based cohort. For this purpose, we assessed genotype and health-related data from 408,354 participants in the UK Biobank. We show that constipation is associated with overall and individual MACE subgroups, and that hypertensive patients with constipation have higher risk of MACE and death, even after adjustment for CCBs and other traditional cardiovascular risk factors. Finally, we detected positive genetic correlations that may explained some of these results.

## Methods

### Study subjects

The UK Biobank (UKBB) is a longitudinal cohort consisting of around 500,000 individuals aged between 40 and 69 years, who were recruited between 2006 and 2010 within the UK.^31^ The data from UKBB includes participants’ genotypes and a wide range of health-related information. This information encompasses electronic medical records (EMR) from hospital inpatient admissions, questionnaire data about general health and lifestyle, as well as self-reported medical conditions and medication usage. After excluding individuals of non-European ancestry, those who withdrew their consent, participants with phenotype/genotype sex mismatches, and those with incomplete or low-quality imputed genetic data, a total of 408,354 participants were included in the analyses (see **Supplementary Table 1**). UKBB follows the principles of the Declaration of Helsinki and received ethical approval from the competent Research Ethics Committee (REC reference for UKBB is 11/NW/0382). The data was obtained under Application Number 86879 from UKBB.

### Phenotype definition

#### Cardiovascular phenotypes

Participants were identified as patients with MACE on a previously reported method,^32^ which incorporated their hospitalization medical records (either primary or secondary), death register data due to ACS, ischemic stroke, or HF, and surgical records related to revascularization procedures, including percutaneous coronary intervention or coronary artery bypass graft. Essential hypertension was identified based on the hospital-inpatient records bearing the ICD10 (International Classification of Diseases, 10^th^ revision) code "I10" section (either primary or secondary) or through self-reported medical history. Additional information on the phenotypic definitions can be found in **Supplementary Table 2**.

#### Constipation

Constipation was identified through medical records with any encounters of ICD10 code "K59.0". The study also incorporated additional constipation phenotypes in sensitivity analyses, including IBS-C and functional constipation, as identified through the ROME III criteria, obtained from the Digestive Health Questionnaire (DHQ) data in the UKBB.^33,34^ Furthermore, participants who routinely consumed laxatives were viewed as a surrogate phenotype for constipation and were, thus, included in the analysis. Further details about these definitions are available in **Supplementary Tables 2 and 3**.

#### Constipating medication usage

Constipation often occurs as a frequent side effect of certain medications, such as a type of commonly used CCBs, an antihypertensive agent. In this study, participants who regularly used CCBs were identified based on their self-reported treatment or medication code, as detailed in **Supplementary Table 2**.

#### Standard modifiable cardiovascular risk factors (SMuRFs)

The occurrence of MACE is recognized to be associated with SMuRFs including hypertension, diabetes, smoking, and hypercholesterolemia.^35^ The definitions of SMuRFs were determined based on inpatient diagnoses from medical records, self-reported medical histories, or the self-reported usage of insulin or cholesterol-lowering medication. Further details are provided in **Supplementary Table 2**.

### Logistic regression association tests

#### Association between MACE and constipation

Initially, the relationship between constipation and the risk of MACE was determined using a logistic regression association test that adjusted for sex, age, and BMI as association analysis Model 1. Sensitivity analyses were then conducted using the following multivariate logistic regression models: (i) adjusting for age, sex, BMI, and the use of CCBs (constipation-inducing medication) as Model 2; (ii) Model 2 with additional adjustments for SMuRFs, including smoking status, hypertension, diabetes, and hypercholesterolemia as Model 3.

#### MACE in hypertensive patients with comorbid constipation

The incidence of MACE in patients with hypertension was assessed by comparing those with comorbid constipation (ICD) to those with normal bowel habits. This comparison was performed using a logistic regression model adjusted for age, sex, BMI, use of CCBs, and SMuRFs.

### Survival analysis

A survival analysis was conducted in hypertensive patients relative to their comorbidity status with constipation. Only patients with MACE, hypertension, and constipation, as defined by ICD codes, were included in this analysis, given that they had valid recorded diagnostic date and time. In addition, in this study, inclusion criteria were restricted to individuals where the initial diagnosis of MACE occurred subsequent to the initial diagnosis of hypertension. The number of survival days was calculated from the date of hypertension diagnosis to the date of the first occurrence of MACE. The risk estimates of MACE in hypertensive patients with constipation, as compared to those with regular bowel habits (reference group), were derived from a Cox proportional hazards regression analysis. This analysis was adjusted for age, sex, BMI, use of CCBs, and SMuRFs.

### Genome-wide association studies of constipation

The genotypic data were sourced from UKBB participants using two different genotyping arrays (UKBB Axiom array and UK BiLEVE Axiom array), with over 95% shared content. The array genotype data underwent stringent quality control procedures before imputation, which was carried out using both the Haplotype Reference Consortium (HRC) and the UK10K + 1000 Genomes Phase 3 reference panels.^31^ Before proceeding with the association analyses, we removed markers with subpar imputation accuracy (INFO score <0.8) or minor allele frequency (MAF) less than 1%.

Genome-wide association analysis (GWAS) of constipation was performed on ICD-diagnosis defined constipation phenotype, using a linear mixed model with BOLT-LMM v2.3.2 including age, sex, BMI, top 10 principal components, genotyping array and CCBs usage.^36^

### Genetic correlation analyses

Publicly accessible GWAS summary statistics for ACS, ischemic stroke, and HF were retrieved from the most recent large-scale GWAS meta-analysis.^37-39^ The SNP heritability (h^2^_SNP_) of individual MACE and constipation phenotypes, along with their genetic correlations, were calculated using LD Score Regression (LDSC) version 1.0.1.^40^

### Data analysis and visualisation tools

All statistical analyses were conducted using R statistical computing environment version 4.1.3.^41^ The association between sex and the prevalence of each phenotype was assessed using Fisher’s exact test. The built-in function glm() was utilized for logistic regression analyses. The "survival" package in R was employed for survival analysis. For visualizations, forest plots and survival curves were generated using the "ggplot2" package, while Manhattan plots for GWAS summary statistics were created using the "qqman" package in R. The "corrplot" package was used to generate genetic correlation plots. Lastly, Venn diagrams were created using the online tool nVenn (http://degradome.uniovi.es/cgi-bin/nVenn/nvenn.cgi).

## Results

### Cohort characteristics

After implementing quality control measures on the sample data, a total of 408,354 participants remained for this study (see **Supplementary Table 1**). Among these, we identified 46,891 (11.5%) individuals who had at least one encounter of MACE (29,704 for ACS, 12,847 for ischemic stroke, and 14,099 for HF, respectively). Using the ICD-10 code K59.0, we detected 23,814 (5.8%) cases of constipation. The demographic and clinical characteristics of these participants are presented in **Table 1**.

**Table 1.**
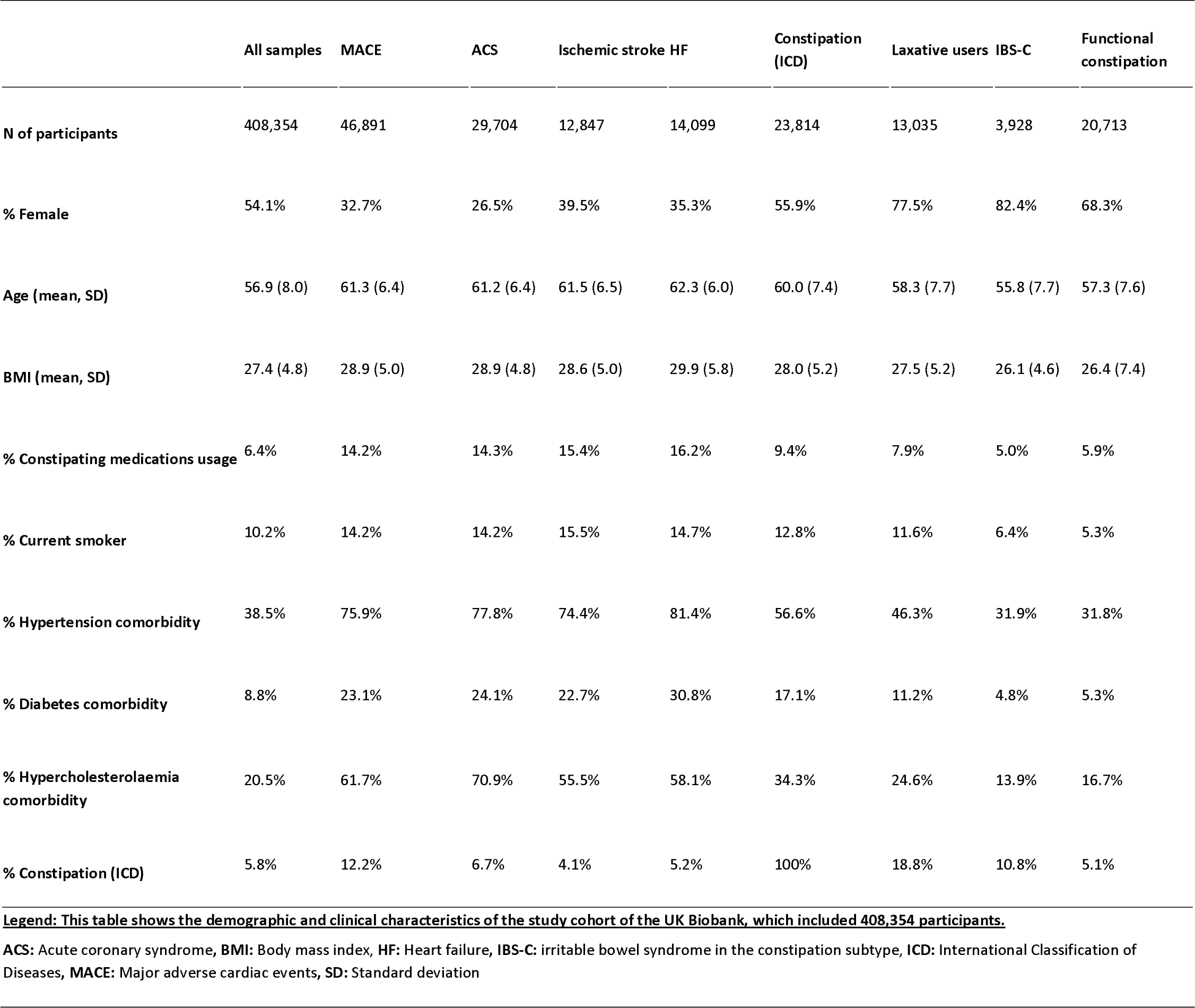
Demographic and clinical characteristics of the study cohort.

It is noteworthy that substantial gender disparities exist in the prevalence of constipation phenotypes, with a higher prevalence observed in females for constipation (ICD) (55.9% females, P<0.05) and other related phenotypes, as detailed in **Table 1**. Conversely, a significantly greater proportion of males was evident in overall MACE (67.3%) and in individual MACE categories (73.5%, 60.5%, and 64.7% for ACS, Stroke, and HF, respectively, P<0.05 for all, **Table 1**). Furthermore, male predominance was also marked in individual SMuRFs, with respective percentages of 53.7%, 52.8%, 60.3%, and 72.0% for smoking status, hypertension, diabetes, and hypercholesterolemia (P<0.05 for all). To account for these observed gender effects, sex was adjusted for in all our association analyses.

The mean (SD) age of both groups was similar: 60.0 (7.4) and 61.3 (6.4) years old, respectively, for cases of constipation and MACE. Among constipated patients, 56.6% presented with hypertension, 12.8% with positive smoking status, 17.1% with diabetes, and 34.3% with hypercholesterolemia. Of note, SMuRFs were more prevalent among individuals with MACE: 75.9% had hypertension, 14.2% were smokers, 23.1% had diabetes, and 61.7% had hypercholesterolemia.

### Associations between Constipation and risk of MACE

Participants with ICD-defined constipation demonstrated a significantly increased risk of MACE compared to those with regular bowel habits (OR=2.15, P<1.00×10^-300^ from a logistic regression analysis adjusted for age, sex, and BMI). Constipation was also significantly associated with individual MACE subgroups: HF (OR=2.72, P<1.00×10^-300^), ischemic stroke (OR=2.36, P=2.02×10^-230^), and ACS (OR=1.62, P=5.82×10^-113^), as reported in **Table 2** and **Figure 1a**.

**Figure 1.**
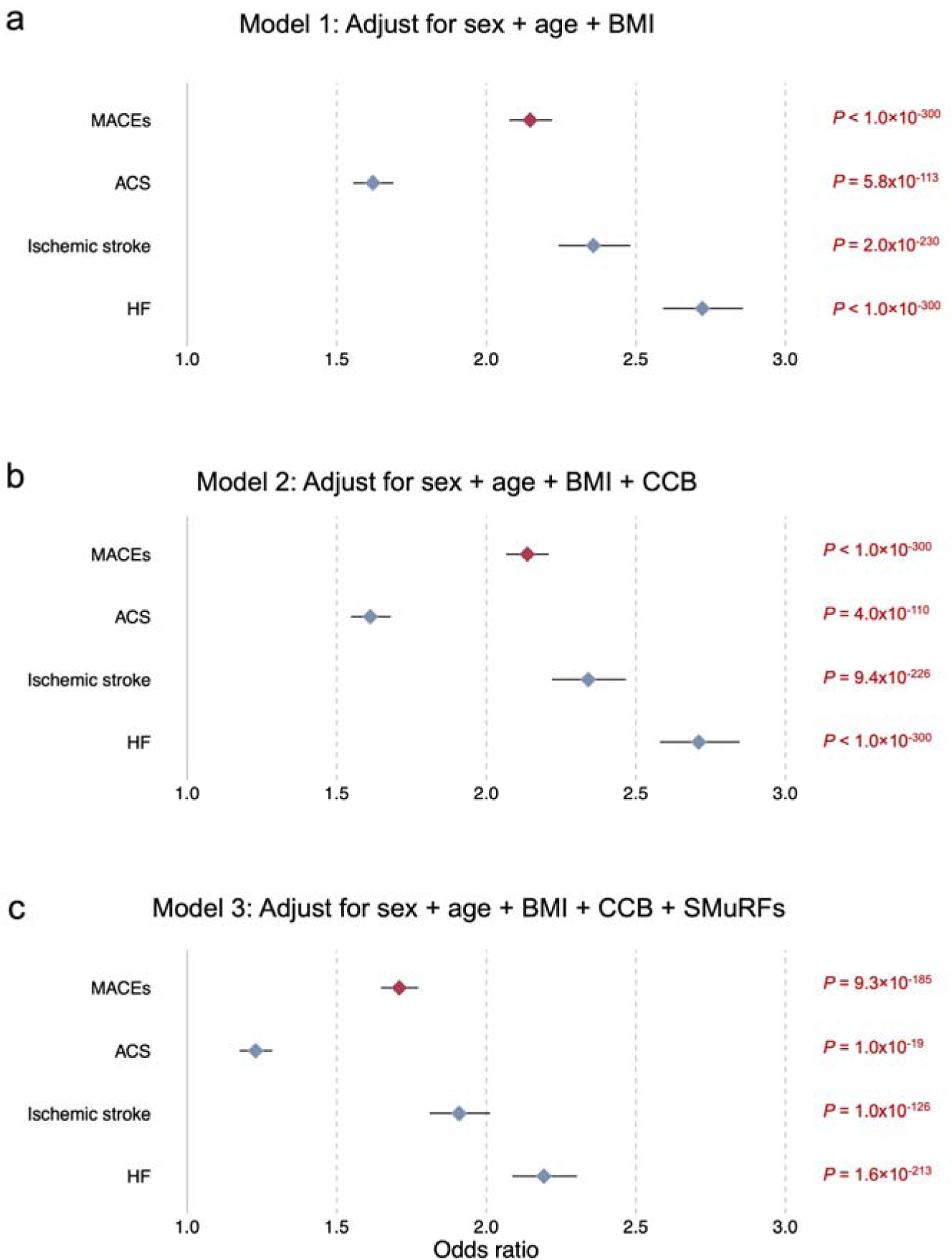
Forest plot showing the association of constipation with the risk of major adverse cardiac events (MACE). These forest plots depict the association between constipation and the risk of MACE, as well as the three individual subgroups within the general population cohort of the UK Biobank, which encompasses 408,354 participants. Within this cohort, we observe a total of 23,814 individuals with Constipation (ICD) and 46,891 participants who experienced MACE (29,704 with Acute Coronary Syndrome (ACS), 12,847 with Stroke, and 14,099 with Heart Failure (HF), respectively). The odds ratios are represented by diamonds, with red indicating MACE and blue signifying the subgroups. The horizontal lines extending from the diamonds represent the 95% confidence intervals of the odds ratios. **a)** association results from Model 1 (adjust for sex, age and BMI), **b)** association results from Model 2 (adjust for sex, age, BMI and use of CCBs), **a)** association results from Model 1 (adjust for sex, age, BMI, use of CCBs and SMuRFs). **ACS**: Acute coronary syndrome, **HF**: Heart failure, **BMI**: Body mass index, **CCBs**: Calcium channel blockers, **SMuRFs**: Standard modifiable cardiovascular risk factors, including hypertension, diabetes, smoking status, and hypercholesterolemia.

**Table 2.** Association test and sensitivity analysis results between MACE and Constipation (based on ICD).

| MACE group | Model 1:<br>Adjusted for sex + age + BMI |  | Model 2:<br>Adjusted for sex + age + BMI +<br>CCB |  | Model 3:<br>Adjusted for sex + age + BMI +<br>CCB + SMuRFs |  |
| --- | --- | --- | --- | --- | --- | --- |
|  | Odds ratio [95% CI] | P-value | Odds ratio [95% CI] | P-value | Odds ratio [95% CI] | P-value |
| MACE | 2.15 [2.08-2.22] | $<1.00 \times 10^{-300}$ | 2.14 [0-2.21] | $<1.00 \times 10^{-300}$ | 1.71 [1.65-1.77] | $9.26 \times 10^{-185}$ |
| ACS | 1.62 [1.56-1.69] | $5.82 \times 10^{-113}$ | 1.61 [0-1.68] | $3.96 \times 10^{-110}$ | 1.23 [1.18-1.28] | $1.02 \times 10^{-19}$ |
| Ischemic stroke | 2.36 [2.24-2.48] | $2.02 \times 10^{-230}$ | 2.34 [0-2.47] | $9.40 \times 10^{-226}$ | 1.91 [1.81-2.01] | $1.03 \times 10^{-126}$ |
| HF | 2.72 [2.59-2.86] | $<1.00 \times 10^{-300}$ | 2.71 [0-2.85] | $<1.00 \times 10^{-300}$ | 2.19 [2.09-2.3] | $1.61 \times 10^{-213}$ |

ACS: Acute coronary syndrome, BMI: Body mass index, CCB: Calcium channel blockers, HF: Heart failure, ICD: International Classification of Diseases, CI: Confidence interval, MACE: Major adverse cardiac events, SMuRFs: Standard modifiable cardiovascular risk factors, including hypertension, diabetes, smoking status, and hypercholesterolemia.

To validate the observed association between constipation and MACE, we conducted two additional sensitivity analyses. Initially, we performed the same regression analyses but also accounted for the use of CCBs, which can impact gut motility (Model 2, see **Methods**). This analysis yielded consistent association results (**Table 2** and **Figure 1b)**. In the second analysis, we included SMuRFs (specifically diabetes, smoking status, hypertension, and hypercholesterolemia) as covariates along with CCBs usage (Model 3, see **Methods**). SMuRFs-adjusted analysis, once again, replicated the significant MACE associations initially observed (P<0.05), despite decreasing the strength of association (OR reduced from 2.15 to 1.71) (**Table 2** and **Figure 1c**).

Lastly, we evaluated the association of MACE with other constipation phenotypes or surrogates. Out of the 408,354 participants, we identified 13,035 who frequently used laxatives. Furthermore, we carried out similar analyses among the 146,488 UK Biobank participants who had completed the DHQ (refer to **Methods**). We compared the prevalence of MACE in participants defined with constipation phenotypes by the ROME III criteria, including IBS-C (N=3,928) and functional constipation (N=20,713) (**Supplementary** Figure 1a). Notably, the four different constipation phenotypes exhibited very minimal overlap among the participants (see **Supplementary** Figure 1b). This analysis revealed that a significantly higher prevalence of MACE was also present in all other three definitions of constipation (see **Supplementary Table 4**). In particular, laxative users showed a stronger association with MACE (OR range=1.40–1.69 across the three adjusted models) than IBS-C (OR range=1.18–1.33) and functional constipation (OR range=1.09– 1.10).

### Risk of MACE in hypertensive patients with comorbid constipation

We identified 157,414 participants with one or more recorded instance of an essential hypertension diagnosis based on the ICD10 code "I10" or through self-reported medical history (see **Supplementary Table 2** for details). Of these, 8.6% (N= 13,469) had a comorbid constipation (ICD) condition. When compared to hypertensive patients with regular bowel habits, those with comorbid constipation showed significantly higher odds of MACE (OR=1.68, P=1.05×10^-136^, from a logistic regression analysis adjusted for age, sex, BMI, use of CCBs, and SMuRFs).

The subset for survival analysis comprised 110,524 participants, all of whom had their first diagnosis of MACE following their initial diagnosis of hypertension. We observed that hypertensive patients with comorbid constipation exhibited a significantly higher risk of experiencing MACE. Compared to hypertensive patients with regular bowel habits (N=99,608), the multivariate-adjusted hazard ratio for patients with constipation (N=10,916) was 1.34 (P=2.3×10^-50^), indicating a 34% increased risk (**Figure 2**). This suggests that constipation may be a significant predictor of MACE in patients with hypertension, even after controlling for other traditional risk factors (see **Methods**).

**Figure 2.**
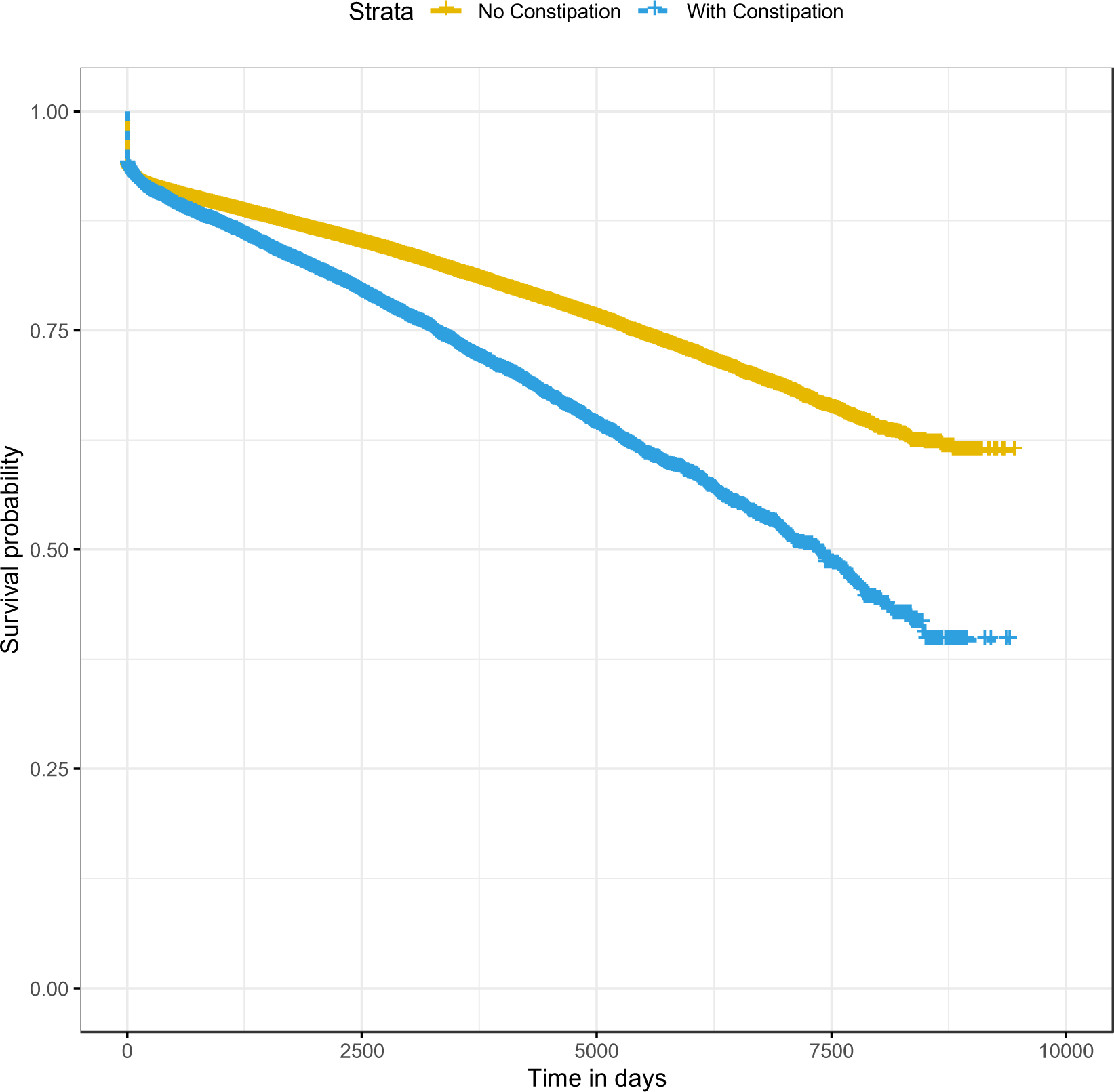
Survival analysis in hypertensive patients with comorbid constipation. This graph illustrates the survival probability over time for hypertensive patients with and without comorbid constipation (ICD) being compared in the study. The x-axis represents time (in days), and the y-axis represents the probability of survival (occurrence of MACE). Yellow line in the plot corresponds to hypertensive patients with normal bowel habits (N=99,608), and blue line to those with comorbid constipation (N=10,916).

### Genetic correlations between constipation and MACE

Quality control procedures on the genetic data yielded a total of 9,572,556 SNP markers for the 408,354 participants in the study. While the GWAS analysis for constipation (ICD) did not yield any signals associated at genome-wide significance (P<5×10^-8^, **Supplementary** Figure 2), the SNP-based heritability (h^2^_SNP_) of constipation was estimated to be around 4% (P=5.9×10^-3^).

We utilized GWAS summary statistics from publicly available meta-analyses for ACS, ischemic stroke, and HF (see **Methods** and **Supplementary Table 5**). Genetic correlation of constipation with these individual MACE subgroups were conducted through LDSC analysis. This revealed a significant genetic overlap of constipation with ACS (r_g_=0.27, P=2.12×10^-6^), ischemic stroke (r_g_=0.23, P=0.011), and HF (r_g_=0.21, P=0.0062) (see **Figure 3**).

**Figure 3.**
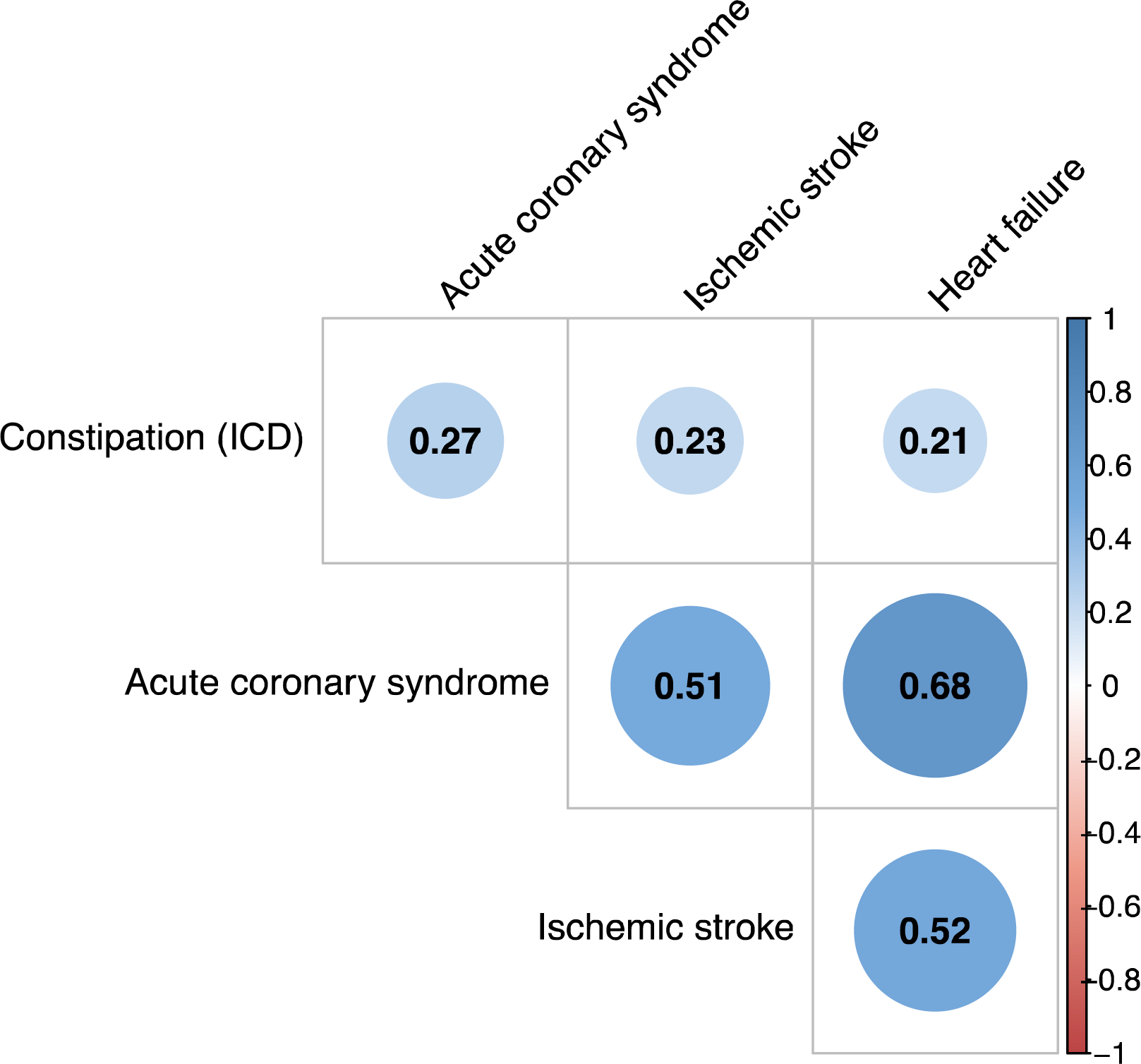
Genetic correlation between constipation and MACE subgroups. This plot provides a visualization of the genetic correlation analysis in this study. In the genetic analysis for constipation, 23,814 participants identified as Constipation (ICD) were compared against 351,891 participants who do not have any form of constipation, serving as study controls. For other publicly available Genome-Wide Association Studies (GWAS), Acute Coronary Syndrome (ACS) involved a comparison between 61,000 cases and 577,000 controls, Ischemic Stroke included 67,162 cases in contrast to 454,450 controls, and Heart Failure (HF) analysis compared 47,309 cases with 930,014 controls (**Supplementary Table 5**). Each cell represents the correlation between the two corresponding traits, with the size and colour intensity of the circles indicating the strength of the correlation. The scale for correlation values (r_g_) is provided in the circle of each cell. Positive correlations are displayed in blue, and negative correlations in red, as demonstrated by the colour scale provided on the right side of the plot. **ICD**: International Classification of Diseases.

## Discussion

In a large-scale survey involving 408,354 individuals in the UK population, we identified constipation as a potential and independent risk factor for MACE. This finding was significant even after controlling for traditional cardiovascular risk factors, including age, sex, BMI, hypertension, diabetes, hypercholesterolemia, smoking status, and the use of anti-hypertensive medication that causes constipation. A two to three times increased risk was detected for HF, ischemic stroke and ACS, in line with previous findings.^9-13^ As a novel finding, our survival analysis demonstrates constipation provides an additional risk for MACE in patients with high blood pressure, the most common modifiable risk factor for death and CVDs.^3^ Remarkably, we show the presence of constipation in hypertensive patients increases the risk of MACE by approximately 1.7 times and contributes to a 34% increase in the risk of death. We also demonstrate this association extends to the genetic level: GWAS and correlation analyses revealed constipation shares up to 27% of genetic risk factors with individual MACE.

These shared genetic factors may underline pathophysiological mechanisms that explain the link between MACE and constipation. This includes disturbances in the autonomic nervous system, which regulates both cardiovascular and GI functions, with the vagus nerve playing a pivotal role.^15,16,42^ Indeed, patients with reduced GI motility often exhibit impaired autonomic vagal function, manifesting in altered heart rate variability and over-activation of the sympathetic nervous system.^43^ Prolonged sympathetic activity stimulation and reduced vagal tone can contribute to elevated blood pressure and heart rate.^15^ As another shared risk factor, dehydration can simultaneously lead to constipation and high blood pressure. Inadequate intake of fluids increase stool viscosity, leading to constipation.^24^ At the same time, hypohydration elevates plasma sodium osmolality, which activates the renin-angiotensin-aldosterone system and raise blood pressure.^25^

Other potential mechanisms linking the gut and the heart may involve alterations in the gut microbiota, known as dysbiosis, disruption of the gut barrier, and subsequent activation of the immune system. Multiple lines of evidence indicate the presence of dysbiosis in constipated and CVD patients.^18,44^ Dysbiosis-associated breakdown of the gut barrier can increase the translocation of microbes and microbial substances (such as metabolites) into the systemic circulation, disrupting intestinal mechanotransduction and triggering inflammatory processes.^45^ Low-grade inflammation is a recognized risk factor for CVD, as it can impair vascular function and contribute to arterial stiffness, leading to hypertension and thrombotic complications.^17,44^ Also important in this context, microbial metabolites, such as short-chain fatty acids (SCFAs) can modulate blood pressure^46-48^ and colonic contractile properties, being altered in both patients with GI dysmotility (including IBS-C) and hypertension.^17,49-51^

Significant association with MACE was observed for all constipation definitions analysed in the current study. Notably, the strength of association was more pronounced in the cohort identified through hospital-inpatient records (based on the ICD K59.0 code) compared to other constipation surrogates and phenotypes, including laxative users, functional constipation, and IBS-C (which involves chronic abdominal pain and/or discomfort).^14,34^ This observation may imply that individuals with hospital-inpatient records related to a constipation episode – and, thus, likely experiencing a more severe form of constipation – face the highest risk of cardiovascular events. Therefore, managing severe constipation through dietary modifications, increased physical activity, adequate hydration, and, if necessary, medications may hold potential to reduce cardiovascular risk profile. This can be performed via increase intake of fibre, which ameliorates GI function^52^ and reduces blood pressure.^53,54^ This warrants further investigations, as it may contribute to alleviating the burden of CVD in the healthcare system.

Our GWAS provides initial evidence of shared genetic predisposing factors between constipation and CVDs. In this regard, constipation-CVD genetic links had been previously reported in relation to ion channels genes, particularly *SCN5A*, coding for the NaV1.5 sodium channel. *SCN5A* plays important roles in the pacemaker activity in the enteric nervous system and in the heart, and variants and loss-of-function variants in this gene have been implicated in cardiac arrythmias and IBS-C.^28,29,55^ Another relevant previous finding is from a GWAS meta-analysis on stool frequency (used as a proxy for gut motility), which identified the rs10407548 locus near the *FFAR3* gene to be associated with individuals reporting less frequent stools.^56^ Interestingly, the *FFAR3* gene codes for the G-protein coupled receptor GPR41, which plays a key role in SCFA signalling and blood pressure regulation.^47,57,58^ *GPR41/43* knockout mice show exaggerated hypertensive and end-organ damage phenotypes, and expression quantitative loci (eQTL) associated with lower GPR41/43 expression are more prevalent in human patients with hypertension.^59^ Hence, altered SCFA-related genes may represent another shared pathogenetic mechanism between constipation and MACE. Genetic links between constipation and CVD may offers significant opportunities to uncover new therapeutic targets for individuals with concomitant CVD and constipation, thus aiding decrease poly-medication and treatment burden. While our current analyses lacked the power to detect loci associated with constipation at genome-wide significance, future larger genomic analyses in more diverse population groups aiming at identifying specific genetic risk factors shared between constipation and MACE will be necessary.

Our study has some limitations. Firstly, despite addressing potential confounding factors linked to traditional cardiovascular risk factors, our analyses lack adjustments for physical activity, dietary habits (e.g., water or fibre intake), and mental health variables. These shared risk factors between constipation and CVD require further in-depth investigation in future studies. Secondly, we only controlled our analysis for the use of CCBs as common antihypertensive agents that can lead to secondary constipation. Although less frequent, we acknowledge that other medications, such as diuretics and beta-blockers, can also cause constipation.^60,61^ Thirdly, due to the limited number of individuals from other populations in the UKBB and aiming for meaningful statistical power, our analyses only included individuals of genetic European-ancestry. Therefore, our findings cannot be generalized to other ancestral populations. Finally, we were unable to thoroughly investigate the directionality of the genetic relationship between constipation and MACE using a mendelian randomization approach, as our constipation GWAS did not detect signals associated at genome-wide significance that could serve as robust genetic instruments.

In conclusion, we identified constipation as a potential risk factor independently associated with higher MACE prevalence, with evidence from both epidemiological and genetic analyses. These findings warrant further studies on their causal relationship and key pathophysiological mechanisms involved in the interactions between constipation and MACE at the molecular level.

## Data Availability

Data used in this study is from the UK Biobank. Access to the data should be requested via the UK Biobank website.

https://www.ukbiobank.ac.uk/

## Acknowledgement

We would like to acknowledge Monash Bioinformatics Platform for access to the M3 servers, and the participants of the UK BioBank.

## References

1. Vaduganathan M, Mensah GA, Turco JV, Fuster V, Roth GA. The Global Burden of Cardiovascular Diseases and Risk: A Compass for Future Health. J Am Coll Cardiol 2022;80:2361–2371.

2. Timmis A, Vardas P, Townsend N, Torbica A, Katus H, De Smedt D, Gale CP, Maggioni AP, Petersen SE, Huculeci R, Kazakiewicz D, de Benito Rubio V, Ignatiuk B, Raisi-Estabragh Z, Pawlak A, Karagiannidis E, Treskes R, Gaita D, Beltrame JF, McConnachie A, Bardinet I, Graham I, Flather M, Elliott P, Mossialos EA, Weidinger F, Achenbach S, Atlas Writing Group ErSoC. European Society of Cardiology: cardiovascular disease statistics 2021. Eur Heart J 2022;43:716-799.

3. Fuchs FD, Whelton PK. High Blood Pressure and Cardiovascular Disease. Hypertension 2020;75:285–292.

4. Lewington S, Clarke R, Qizilbash N, Peto R, Collins R, Collaboration PS. Age-specific relevance of usual blood pressure to vascular mortality: a meta-analysis of individual data for one million adults in 61 prospective studies. Lancet 2002;360:1903–1913.

5. Pencina MJ, Navar AM, Wojdyla D, Sanchez RJ, Khan I, Elassal J, D’Agostino RB, Peterson ED, Sniderman AD. Quantifying Importance of Major Risk Factors for Coronary Heart Disease. Circulation 2019;139:1603–1611.

6. Yusuf S, Hawken S, Ounpuu S, Dans T, Avezum A, Lanas F, McQueen M, Budaj A, Pais P, Varigos J, Lisheng L, Investigators IS. Effect of potentially modifiable risk factors associated with myocardial infarction in 52 countries (the INTERHEART study): case-control study. Lancet 2004;364:937–952.

7. O’Donnell JA, Zheng T, Meric G, Marques FZ. The gut microbiome and hypertension. Nat Rev Nephrol 2023;19:153–167.

8. Peh A, O’Donnell JA, Broughton BRS, Marques FZ. Gut Microbiota and Their Metabolites in Stroke: A Double-Edged Sword. Stroke 2022;53:1788–1801.

9. Sundbøll J, Szépligeti SK, Adelborg K, Szentkúti P, Gregersen H, Sørensen HT. Constipation and risk of cardiovascular diseases: a Danish population-based matched cohort study. BMJ Open 2020;10:e037080.

10. Judkins CP, Wang Y, Jelinic M, Bobik A, Vinh A, Sobey CG, Drummond GR. Association of constipation with increased risk of hypertension and cardiovascular events in elderly Australian patients. Sci Rep 2023;13:10943.

11. Salmoirago-Blotcher E, Crawford S, Jackson E, Ockene J, Ockene I. Constipation and risk of cardiovascular disease among postmenopausal women. Am J Med 2011;124:714–723.

12. Sumida K, Molnar MZ, Potukuchi PK, Thomas F, Lu JL, Yamagata K, Kalantar-Zadeh K, Kovesdy CP. Constipation and risk of death and cardiovascular events. Atherosclerosis 2019;281:114–120.

13. Ishiyama Y, Hoshide S, Mizuno H, Kario K. Constipation-induced pressor effects as triggers for cardiovascular events. J Clin Hypertens (Greenwich) 2019;21:421–425.

14. Scott SM, Simrén M, Farmer AD, Dinning PG, Carrington EV, Benninga MA, Burgell RE, Dimidi E, Fikree A, Ford AC, Fox M, Hoad CL, Knowles CH, Krogh K, Nugent K, Remes-Troche JM, Whelan K, Corsetti M. Chronic constipation in adults: Contemporary perspectives and clinical challenges. 1: Epidemiology, diagnosis, clinical associations, pathophysiology and investigation. Neurogastroenterol Motil 2021;33:e14050.

15. Capilupi MJ, Kerath SM, Becker LB. Vagus Nerve Stimulation and the Cardiovascular System. Cold Spring Harb Perspect Med 2020;10.

16. Bonaz B, Sinniger V, Pellissier S. Vagal tone: effects on sensitivity, motility, and inflammation. Neurogastroenterol Motil 2016;28:455–462.

17. O’Donnell JA, Zheng T, Meric G, Marques FZ. The gut microbiome and hypertension. Nat Rev Nephrol 2023.

18. Ohkusa T, Koido S, Nishikawa Y, Sato N. Gut Microbiota and Chronic Constipation: A Review and Update. Front Med (Lausanne) 2019;6:19.

19. Lai H, Li Y, He Y, Chen F, Mi B, Li J, Xie J, Ma G, Yang J, Xu K, Liao X, Yin Y, Liang J, Kong L, Wang X, Li Z, Shen Y, Dang S, Zhang L, Wu Q, Zeng L, Shi L, Zhang X, Tian T, Liu X. Effects of dietary fibers or probiotics on functional constipation symptoms and roles of gut microbiota: a double-blinded randomized placebo trial. Gut Microbes 2023;15:2197837.

20. van der Schoot A, Drysdale C, Whelan K, Dimidi E. The Effect of Fiber Supplementation on Chronic Constipation in Adults: An Updated Systematic Review and Meta-Analysis of Randomized Controlled Trials. Am J Clin Nutr 2022;116:953–969.

21. Kaye DM, Shihata WA, Jama HA, Tsyganov K, Ziemann M, Kiriazis H, Horlock D, Vijay A, Giam B, Vinh A, Johnson C, Fiedler A, Donner D, Snelson M, Coughlan MT, Phillips S, Du XJ, El-Osta A, Drummond G, Lambert GW, Spector TD, Valdes AM, Mackay CR, Marques FZ. Deficiency of Prebiotic Fiber and Insufficient Signaling Through Gut Metabolite-Sensing Receptors Leads to Cardiovascular Disease. Circulation 2020;141:1393–1403.

22. Iovino P, Chiarioni G, Bilancio G, Cirillo M, Mekjavic IB, Pisot R, Ciacci C. New onset of constipation during long-term physical inactivity: a proof-of-concept study on the immobility-induced bowel changes. PLoS One 2013;8:e72608.

23. Zhang X, Cash RE, Bower JK, Focht BC, Paskett ED. Physical activity and risk of cardiovascular disease by weight status among U.S adults. PLoS One 2020;15:e0232893.

24. Arnaud MJ. Mild dehydration: a risk factor of constipation? Eur J Clin Nutr 2003;57 Suppl 2:S88-95.

25. Watso JC, Farquhar WB. Hydration Status and Cardiovascular Function. Nutrients 2019;11.

26. Krevsky B, Maurer AH, Niewiarowski T, Cohen S. Effect of verapamil on human intestinal transit. Dig Dis Sci 1992;37:919–924.

27. Castell DO. Calcium-channel blocking agents for gastrointestinal disorders. Am J Cardiol 1985;55:210B–213B.

28. Darbar D, Kannankeril PJ, Donahue BS, Kucera G, Stubblefield T, Haines JL, George AL, Roden DM. Cardiac sodium channel (SCN5A) variants associated with atrial fibrillation. Circulation 2008;117:1927–1935.

29. Beyder A, Mazzone A, Strege PR, Tester DJ, Saito YA, Bernard CE, Enders FT, Ek WE, Schmidt PT, Dlugosz A, Lindberg G, Karling P, Ohlsson B, Gazouli M, Nardone G, Cuomo R, Usai-Satta P, Galeazzi F, Neri M, Portincasa P, Bellini M, Barbara G, Camilleri M, Locke GR, Talley NJ, D’Amato M, Ackerman MJ, Farrugia G. Loss-of-function of the voltage-gated sodium channel NaV1.5 (channelopathies) in patients with irritable bowel syndrome. Gastroenterology 2014;146:1659–1668.

30. Strege PR, Mazzone A, Bernard CE, Neshatian L, Gibbons SJ, Saito YA, Tester DJ, Calvert ML, Mayer EA, Chang L, Ackerman MJ, Beyder A, Farrugia G. Irritable bowel syndrome patients have SCN5A channelopathies that lead to decreased NaV1.5 current and mechanosensitivity. Am J Physiol Gastrointest Liver Physiol 2018;314:G494–G503.

31. Bycroft C, Freeman C, Petkova D, Band G, Elliott LT, Sharp K, Motyer A, Vukcevic D, Delaneau O, O’Connell J, Cortes A, Welsh S, Young A, Effingham M, McVean G, Leslie S, Allen N, Donnelly P, Marchini J. The UK Biobank resource with deep phenotyping and genomic data. Nature 2018;562:203–209.

32. Choi BG, Rha SW, Yoon SG, Choi CU, Lee MW, Kim SW. Association of Major Adverse Cardiac Events up to 5 Years in Patients With Chest Pain Without Significant Coronary Artery Disease in the Korean Population. J Am Heart Assoc 2019;8:e010541.

33. Eijsbouts C, Zheng T, Kennedy NA, Bonfiglio F, Anderson CA, Moutsianas L, Holliday J, Shi J, Shringarpure S, andMe Research T, Voda AI, Bellygenes I, Farrugia G, Franke A, Hubenthal M, Abecasis G, Zawistowski M, Skogholt AH, Ness-Jensen E, Hveem K, Esko T, Teder-Laving M, Zhernakova A, Camilleri M, Boeckxstaens G, Whorwell PJ, Spiller R, McVean G, D’Amato M, Jostins L, Parkes M. Genome-wide analysis of 53,400 people with irritable bowel syndrome highlights shared genetic pathways with mood and anxiety disorders. Nat Genet 2021;53:1543–1552.

34. Longstreth GF, Thompson WG, Chey WD, Houghton LA, Mearin F, Spiller RC. Functional bowel disorders. Gastroenterology 2006;130:1480-1491.

35. Figtree GA, Vernon ST, Hadziosmanovic N, Sundstrom J, Alfredsson J, Arnott C, Delatour V, Leosdottir M, Hagstrom E. Mortality in STEMI patients without standard modifiable risk factors: a sex-disaggregated analysis of SWEDEHEART registry data. Lancet 2021;397:1085–1094.

36. Loh PR, Tucker G, Bulik-Sullivan BK, Vilhjalmsson BJ, Finucane HK, Salem RM, Chasman DI, Ridker PM, Neale BM, Berger B, Patterson N, Price AL. Efficient Bayesian mixed-model analysis increases association power in large cohorts. Nat Genet 2015;47:284–290.

37. Shah S, Henry A, Roselli C, Lin H, Sveinbjornsson G, Fatemifar G, Hedman AK, Wilk JB, Morley MP, Chaffin MD, Helgadottir A, Verweij N, Dehghan A, Almgren P, Andersson C, Aragam KG, Arnlov J, Backman JD, Biggs ML, Bloom HL, Brandimarto J, Brown MR, Buckbinder L, Carey DJ, Chasman DI, Chen X, Chen X, Chung J, Chutkow W, Cook JP, Delgado GE, Denaxas S, Doney AS, Dorr M, Dudley SC, Dunn ME, Engstrom G, Esko T, Felix SB, Finan C, Ford I, Ghanbari M, Ghasemi S, Giedraitis V, Giulianini F, Gottdiener JS, Gross S, Guethbjartsson DF, Gutmann R, Haggerty CM, van der Harst P, Hyde CL, Ingelsson E, Jukema JW, Kavousi M, Khaw KT, Kleber ME, Kober L, Koekemoer A, Langenberg C, Lind L, Lindgren CM, London B, Lotta LA, Lovering RC, Luan J, Magnusson P, Mahajan A, Margulies KB, Marz W, Melander O, Mordi IR, Morgan T, Morris AD, Morris AP, Morrison AC, Nagle MW, Nelson CP, Niessner A, Niiranen T, O’Donoghue ML, Owens AT, Palmer CNA, Parry HM, Perola M, Portilla-Fernandez E, Psaty BM, Regeneron Genetics C, Rice KM, Ridker PM, Romaine SPR, Rotter JI, Salo P, Salomaa V, van Setten J, Shalaby AA, Smelser DT, Smith NL, Stender S, Stott DJ, Svensson P, Tammesoo ML, Taylor KD, Teder-Laving M, Teumer A, Thorgeirsson G, Thorsteinsdottir U, Torp-Pedersen C, Trompet S, Tyl B, Uitterlinden AG, Veluchamy A, Volker U, Voors AA, Wang X, Wareham NJ, Waterworth D, Weeke PE, Weiss R, Wiggins KL, Xing H, Yerges-Armstrong LM, Yu B, Zannad F, Zhao JH, Hemingway H, Samani NJ, McMurray JJV, Yang J, Visscher PM, Newton-Cheh C, Malarstig A, Holm H, Lubitz SA, Sattar N, Holmes MV, Cappola TP, Asselbergs FW, Hingorani AD, Kuchenbaecker K, Ellinor PT, Lang CC, Stefansson K, Smith JG, Vasan RS, Swerdlow DI, Lumbers RT. Genome-wide association and Mendelian randomisation analysis provide insights into the pathogenesis of heart failure. Nat Commun 2020;11:163.

38. Malik R, Chauhan G, Traylor M, Sargurupremraj M, Okada Y, Mishra A, Rutten-Jacobs L, Giese AK, van der Laan SW, Gretarsdottir S, Anderson CD, Chong M, Adams HHH, Ago T, Almgren P, Amouyel P, Ay H, Bartz TM, Benavente OR, Bevan S, Boncoraglio GB, Brown RD, Jr., Butterworth AS, Carrera C, Carty CL, Chasman DI, Chen WM, Cole JW, Correa A, Cotlarciuc I, Cruchaga C, Danesh J, de Bakker PIW, DeStefano AL, den Hoed M, Duan Q, Engelter ST, Falcone GJ, Gottesman RF, Grewal RP, Gudnason V, Gustafsson S, Haessler J, Harris TB, Hassan A, Havulinna AS, Heckbert SR, Holliday EG, Howard G, Hsu FC, Hyacinth HI, Ikram MA, Ingelsson E, Irvin MR, Jian X, Jimenez-Conde J, Johnson JA, Jukema JW, Kanai M, Keene KL, Kissela BM, Kleindorfer DO, Kooperberg C, Kubo M, Lange LA, Langefeld CD, Langenberg C, Launer LJ, Lee JM, Lemmens R, Leys D, Lewis CM, Lin WY, Lindgren AG, Lorentzen E, Magnusson PK, Maguire J, Manichaikul A, McArdle PF, Meschia JF, Mitchell BD, Mosley TH, Nalls MA, Ninomiya T, O’Donnell MJ, Psaty BM, Pulit SL, Rannikmae K, Reiner AP, Rexrode KM, Rice K, Rich SS, Ridker PM, Rost NS, Rothwell PM, Rotter JI, Rundek T, Sacco RL, Sakaue S, Sale MM, Salomaa V, Sapkota BR, Schmidt R, Schmidt CO, Schminke U, Sharma P, Slowik A, Sudlow CLM, Tanislav C, Tatlisumak T, Taylor KD, Thijs VNS, Thorleifsson G, Thorsteinsdottir U, Tiedt S, Trompet S, Tzourio C, van Duijn CM, Walters M, Wareham NJ, Wassertheil-Smoller S, Wilson JG, Wiggins KL, Yang Q, Yusuf S, Consortium AF, Cohorts for H, Aging Research in Genomic Epidemiology C, International Genomics of Blood Pressure C, Consortium, Starnet, Bis JC, Pastinen T, Ruusalepp A, Schadt EE, Koplev S, Bjorkegren JLM, Codoni V, Civelek M, Smith NL, Tregouet DA, Christophersen IE, Roselli C, Lubitz SA, Ellinor PT, Tai ES, Kooner JS, Kato N, He J, van der Harst P, Elliott P, Chambers JC, Takeuchi F, Johnson AD, BioBank Japan Cooperative Hospital G, Consortium C, Consortium E-C, Consortium EP-I, International Stroke Genetics C, Consortium M, Neurology Working Group of the CC, Network NSG, Study UKYLD, Consortium M, Sanghera DK, Melander O, Jern C, Strbian D, Fernandez-Cadenas I, Longstreth WT, Jr., Rolfs A, Hata J, Woo D, Rosand J, Pare G, Hopewell JC, Saleheen D, Stefansson K, Worrall BB, Kittner SJ, Seshadri S, Fornage M, Markus HS, Howson JMM, Kamatani Y, Debette S, Dichgans M. Multiancestry genome-wide association study of 520,000 subjects identifies 32 loci associated with stroke and stroke subtypes. Nat Genet 2018;50:524-537.

39. Hartiala JA, Han Y, Jia Q, Hilser JR, Huang P, Gukasyan J, Schwartzman WS, Cai Z, Biswas S, Tregouet DA, Smith NL, Consortium I, Group CCHW, Consortium G-C, Seldin M, Pan C, Mehrabian M, Lusis AJ, Bazeley P, Sun YV, Liu C, Quyyumi AA, Scholz M, Thiery J, Delgado GE, Kleber ME, Marz W, Howe LJ, Asselbergs FW, van Vugt M, Vlachojannis GJ, Patel RS, Lyytikainen LP, Kahonen M, Lehtimaki T, Nieminen TVM, Kuukasjarvi P, Laurikka JO, Chang X, Heng CK, Jiang R, Kraus WE, Hauser ER, Ferguson JF, Reilly MP, Ito K, Koyama S, Kamatani Y, Komuro I, Biobank J, Stolze LK, Romanoski CE, Khan MD, Turner AW, Miller CL, Aherrahrou R, Civelek M, Ma L, Bjorkegren JLM, Kumar SR, Tang WHW, Hazen SL, Allayee H. Genome-wide analysis identifies novel susceptibility loci for myocardial infarction. Eur Heart J 2021;42:919–933.

40. Bulik-Sullivan BK, Loh PR, Finucane HK, Ripke S, Yang J, Schizophrenia Working Group of the Psychiatric Genomics C, Patterson N, Daly MJ, Price AL, Neale BM. LD Score regression distinguishes confounding from polygenicity in genome-wide association studies. Nat Genet 2015;47:291–295.

41. Team RC. R: A Language and Environment for Statistical Computing. Vienna, Austria: R Foundation for Statistical Computing, 2022.

42. Yuan Y, Ali MK, Mathewson KJ, Sharma K, Faiyaz M, Tan W, Parsons SP, Zhang KK, Milkova N, Liu L, Ratcliffe E, Armstrong D, Schmidt LA, Chen JH, Huizinga JD. Associations Between Colonic Motor Patterns and Autonomic Nervous System Activity Assessed by High-Resolution Manometry and Concurrent Heart Rate Variability. Front Neurosci 2019;13:1447.

43. Mazurak N, Seredyuk N, Sauer H, Teufel M, Enck P. Heart rate variability in the irritable bowel syndrome: a review of the literature. Neurogastroenterol Motil 2012;24:206–216.

44. Witkowski M, Weeks TL, Hazen SL. Gut Microbiota and Cardiovascular Disease. Circ Res 2020;127:553–570.

45. Lewis CV, Taylor WR. Intestinal barrier dysfunction as a therapeutic target for cardiovascular disease. Am J Physiol Heart Circ Physiol 2020;319:H1227–H1233.

46. Jama HA, Rhys-Jones D, Nakai M, Yao CK, Climie RE, Sata Y, Anderson D, Creek DJ, Head GA, Kaye DM, Mackay CR, Muir J, Marques FZ. Prebiotic intervention with HAMSAB in untreated essential hypertensive patients assessed in a phase II randomized trial. Nature Cardiovascular Research 2023;2:35–43.

47. Kaye DM, Shihata W, Jama HA, Tsyganov K, Ziemann M, Kiriazis H, Horlock D, Vijay A, Giam B, Vinh A, Johnson C, Fiedler A, Donner D, Snelson M, Coughlan MT, Phillips S, Du XJ, El-Osta A, Drummond G, Lambert GW, Spector T, Valdes AM, Mackay CR, Marques FZ. Deficiency of Prebiotic Fibre and Insufficient Signalling Through Gut Metabolite Sensing Receptors Leads to Cardiovascular Disease. Circulation 2020;141:1393–1403.

48. Marques FZ, Nelson E, Chu PY, Horlock D, Fiedler A, Ziemann M, Tan JK, Kuruppu S, Rajapakse NW, El-Osta A, Mackay CR, Kaye DM. High-Fiber Diet and Acetate Supplementation Change the Gut Microbiota and Prevent the Development of Hypertension and Heart Failure in Hypertensive Mice. Circulation 2017;135:964–977.

49. Jiang W, Wu J, Zhu S, Xin L, Yu C, Shen Z. The Role of Short Chain Fatty Acids in Irritable Bowel Syndrome. J Neurogastroenterol Motil 2022;28:540–548.

50. Zheng Z, Tang J, Hu Y, Zhang W. Role of gut microbiota-derived signals in the regulation of gastrointestinal motility. Front Med (Lausanne) 2022;9:961703.

51. Marques FZ, Mackay CR, Kaye DM. Beyond gut feelings: how the gut microbiota regulates blood pressure. Nat Rev Cardiol 2018;15:20–32.

52. Gill SK, Rossi M, Bajka B, Whelan K. Dietary fibre in gastrointestinal health and disease. Nat Rev Gastroenterol Hepatol 2021;18:101-116.

53. Reynolds AN, Akerman A, Kumar S, Diep Pham HT, Coffey S, Mann J. Dietary fibre in hypertension and cardiovascular disease management: systematic review and meta-analyses. BMC Med 2022;20:139.

54. Reynolds A, Mann J, Cummings J, Winter N, Mete E, Te Morenga L. Carbohydrate quality and human health: a series of systematic reviews and meta-analyses. Lancet 2019;393:434–445.

55. Savio-Galimberti E, Darbar D. Atrial Fibrillation and SCN5A Variants. Card Electrophysiol Clin 2014;6:741–748.

56. Bonfiglio F, Liu X, Smillie C, Pandit A, Kurilshikov A, Bacigalupe R, Zheng T, Nim H, Garcia-Etxebarria K, Bujanda L, Andreasson A, Agreus L, Walter S, Abecasis G, Eijsbouts C, Jostins L, Parkes M, Hughes DA, Timpson N, Raes J, Franke A, Kennedy NA, Regev A, Zhernakova A, Simren M, Camilleri M, D’Amato M. GWAS of stool frequency provides insights into gastrointestinal motility and irritable bowel syndrome. Cell Genom 2021;1:None.

57. Onyszkiewicz M, Gawrys-Kopczynska M, Konopelski P, Aleksandrowicz M, Sawicka A, Koźniewska E, Samborowska E, Ufnal M. Butyric acid, a gut bacteria metabolite, lowers arterial blood pressure via colon-vagus nerve signaling and GPR41/43 receptors. Pflugers Arch 2019;471:1441–1453.

58. Natarajan N, Hori D, Flavahan S, Steppan J, Flavahan NA, Berkowitz DE, Pluznick JL. Microbial short chain fatty acid metabolites lower blood pressure via endothelial G protein-coupled receptor 41. Physiol Genomics 2016;48:826–834.

59. Rikeish RM, Tenghao Z, Evany D, Liang X, Anastasia B-W, Hamdi AJ, Michael N, Madeleine P, Chad J, Ekaterina S, Natalie B, Maria-Kaparakis L, David MK, Joanne AOD, Charles RM, Francine ZM. GPR41/43 regulates blood pressure by improving gut epithelial barrier integrity to prevent TLR4 activation and renal inflammation. bioRxiv 2023:2023.2003.2020.533376.

60. Ueki T, Nakashima M. Relationship Between Constipation and Medication. J UOEH 2019;41:145–151.

61. Turkoski BB. "I Can’t Poop": Medication-Induced Constipation. Orthop Nurs 2018;37:192–196.

