## Supplementary material for "Constipation is associated with an increased risk of major adverse cardiac events in a UK population": Online supplemental tables and figures

**Running title**: Constipation and major adverse cardiac events

Tenghao Zheng MD, PhD[1]; Leticia Camargo Tavares, MSc[1]; Mauro D'Amato, PhD[2,3,4]; Francine Z. Marques, PhD[1,5,6*]

[1]Hypertension Research Laboratory, School of Biological Sciences, Faculty of Science, Monash University, Melbourne, Australia; [2]Gastrointestinal Genetics Lab, CIC bioGUNE - BRTA, Derio, Spain; [3]Ikerbasque, Basque Foundation for Science, Bilbao, Spain; [4]Department of Medicine and Surgery, LUM University, Casamassima, Italy; [5]Baker Heart and Diabetes Institute, Melbourne, Australia; [6]Victorian Heart Institute, Monash University, Melbourne, Australia.

***Corresponding author**: A/Prof Francine Marques, Hypertension Research Laboratory, School of Biological Sciences, Faculty of Science, Monash University, Melbourne, Australia, Phone: +61-03-9905 6958.

### Supplementary Figures

A

| | Definition | N of cases |
|---|---|---|
| Constipation (ICD) | ICD10 code K59.0, both primary and secondary | 23,814 |
| Laxatives users | Self-reported laxatives usage | 13,035 |
| IBS-C | Defined according to ROME III criteria in digestive health questionnaire | 3,928 |
| Functional constipation | Defined according to ROME III criteria in digestive health questionnaire | 20,713 |

B

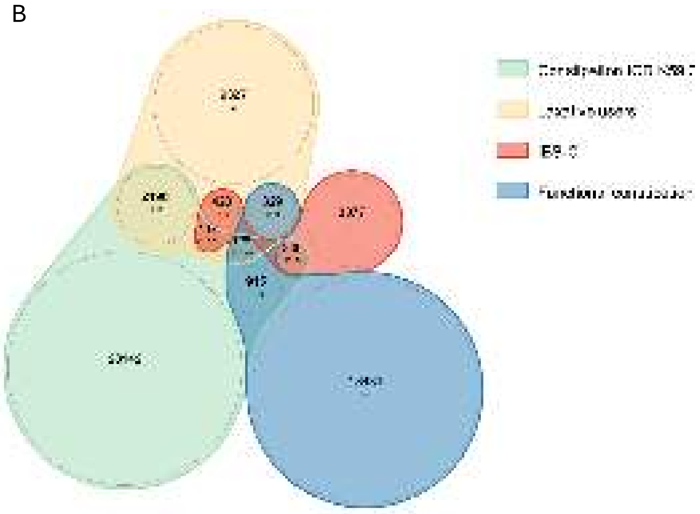

**Supplementary Figure 1.** Overview of the four constipation phenotypes investigated in the study.

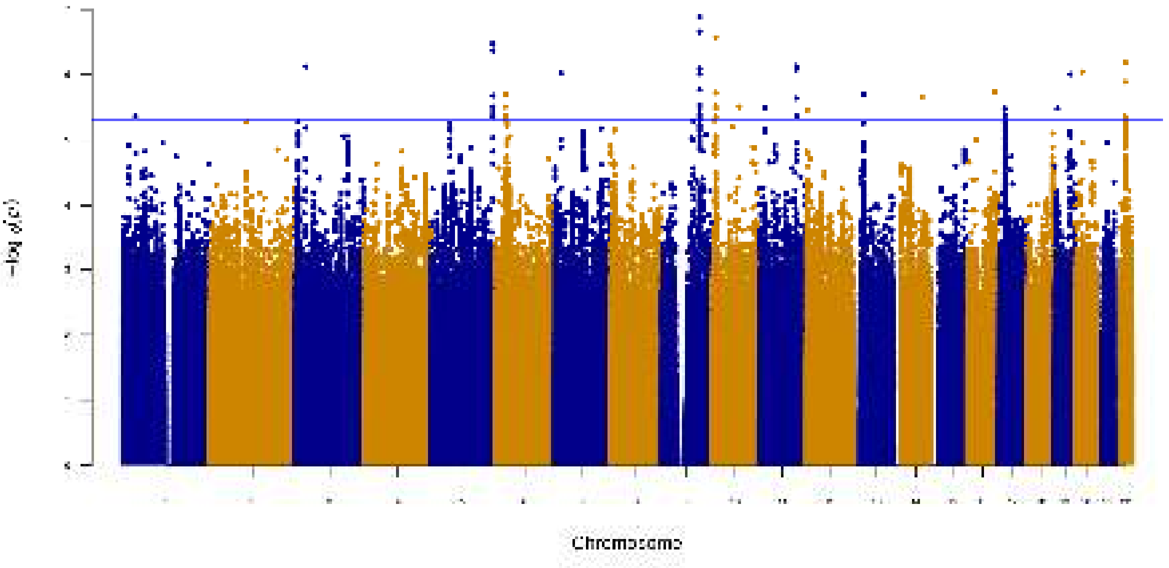


**Supplementary Figure 2.** Manhattan plots of constipation (ICD) GWAS results.

#### Supplementary Tables

**Supplementary Table 1.** Quality control summary of samples in the study cohort.

| Quality control steps | Samples removed | Samples left |
|---|---|---|
| Initial number | 0 | 502,411 |
| Self-report White individuals (British, Irish and other white) | -42,274 | 460,137 |
| Genetic Caucasians | -50,586 | 409,551 |
| Sex mismatch check | -307 | 409,244 |
| Genotype quality check | -727 | 408,517 |
| Remove individuals with excess relatives | -163 | 408,354 |

**Supplementary Table 2.** Phenotype definitions and their associated coding in UK Biobank.

| Category | Phenotype | UK Biobank Data Fields & Coding | | | | | | |
|---|---|---|---|---|---|---|---|---|
| | | **Hospital Inpatient Diagnosis (ICD 10)** | **Self-reported Medical History** | **Surgical code (OPCS 4)** | **Self-reported Surgical History** | **Death register** | **Digestive Health Questionnaire** | **Medication Usage** |
| Constipation | Constipation (ICD) | 41270 = K59.0 | | | | | | |
| | Laxatives users | | | | | | | 6154 = 6 or 20003 = 1140879476 |
| | IBS-C | | | | | | Details in **Supplementary Table 3** | |
| | Functional constipation | | | | | | Details in **Supplementary Table 3** | |
| MACE | Acute coronary syndrome | 41270 in (I20.0, I21.*, I24.8, I24.9) | 20002 = 1075 | 41272 in (K40.*, K41.*, K42.*, K43.*, K44.*, K45.*, K46.*, K49.*, K50.*, K75.*) | 20004 in (1070, 1095) | 40001 in (I20.0, I21.*, I24.8, I24.9) or 40002 in (I20.0, I21.*, I24.8, I24.9) | | |
| | Ischemic stroke | 41270 in (I63.*, I64.*) | 20002 in (1081, 1583) | | | 40001 in (I63.*, I64.*) or 40002 in (I63.*, I64.*) | | |

| | | | | | | |
|---|---|---|---|---|---|---|
| | Heart failure | 41270 in (I50.*) | 20002 = 1076 | | 40001 in (I50.*) or 40002 in (I50.*) | |
| Medication | Constipating medication users (Calcium channel blocker) | | | | | 20003 in (1140879802, 1140879806, 1140926778, 1140888510, 1141153328, 1140879810, 1140888646, 1141165470, 1140861088, 1140860426, 1140861190, 1140928226) |
| SMuRFs | Essential Hypertension | 41270 in (I10.*) | 20002 in (1065, 1072) | | | |
| | Diabetes | 41270 in (E10.*, E11.*, E12.*, E13.*, E14.*) | 20002 in (1220, 1221, 1222, 1223) | | | 6177 = 1 |
| | Hypercholesterolaemia | 41270 = K78.0 | | | | 6177 = 3 |
| | Smoking | | 20116 = 2 | | | |

**Legend:** **MACE:** Major adverse cardiac event; **ICD:** International Classification of Diseases; **OPCS:** Operating Procedure Codes Supplement; **IBS-C:** Irritable bowel syndrome constipation subtype.

**Supplementary Table 3.** Constipation phenotypes defined according to the ROME-III criteria in UK Biobank.

| Criteria | Questions in Digestive Health Questionnaire | Definition of Positive Answer | Definition of IBS-C UK Biobank coding | Definition of Functional constipation UK Biobank coding |
|---|---|---|---|---|
| Criteria 1 | In the last 3 months, how often did you have discomfort or pain anywhere in your abdomen? | 2-3 times per month or more | 21025 in (3,4,5,6) | |
| Criteria 2 | For women: Did this discomfort or pain occur only during your menstrual bleeding and not at other times? | No or not applicable | 21026 in (1,NA) | |
| Criteria 3 | Have you had this discomfort or pain 6 months or longer? | Yes | 21027 = 1 | |
| | How often did this discomfort or pain get better or stop after you had a bowel movement? | At least sometimes | 21028 in (-501,-502,-503,-504) | |
| Criteria 4 (Positive = at least two of the three items) | When this discomfort or pain started, did you have more frequent bowel movements? OR When this discomfort or pain started, did you have less frequent bowel movements? | At least sometimes to at least one of these questions. | 21029 in (-501, -502, -503, -504) or 21030 in (-501, -502, -503, -504) | |
| | When this discomfort or pain started, were your stools (bowel movements) looser? OR When this discomfort or pain started, were your stools (bowel movements) harder? | At least sometimes to at least one of these questions. | 21031 in (-501, -502, -503, -504) or 21032 in (-501, -502, -503, -504) | |

| | | | |
|---|---|---|---|
| Criteria 5 | Frequency of hard/lumpy stools in the last 3 months or Frequency of loose/mushy/watery stools in the last 3 months | At least sometimes for hard/lumpy stools and no loose/mushy/watery stools | Positive for Criteria 1-4 and 21033 in (-501,-502,-503,-504) and 21034 = 0 |

**Legend**: IBS-C: Irritable bowel syndrome constipation subtype; NA: Not applicable.

**Supplementary Table 4.** Association analyses results between MACE and other constipation phenotypes.

| Constipation phenotype | MACE group | Model 1: Adjust for sex + age + BMI | | Model 2: Adjust for sex + age + BMI +CCB | | Model 3: Adjust for sex + age + BMI +CCB + SMuRFs | |
|---|---|---|---|---|---|---|---|
| | | Odds ratio [95% CI] | P | Odds ratio [95% CI] | P | Odds ratio [95% CI] | P |
| Laxatives users | MACEs | 1.69 [1.61-1.78] | 5.32E-88 | 1.67 [1.59-1.76] | 2.00E-84 | 1.4 [1.32-1.48] | 7.55E-33 |
| | ACS | 1.66 [1.55-1.76] | 1.17E-54 | 1.64 [1.54-1.75] | 1.91E-52 | 1.33 [1.24-1.42] | 1.64E-16 |
| | Ischemic stroke | 1.72 [1.58-1.87] | 1.30E-37 | 1.7 [1.56-1.84] | 8.39E-36 | 1.46 [1.34-1.58] | 1.72E-18 |
| | HF | 1.6 [1.47-1.73] | 3.51E-28 | 1.58 [1.45-1.71] | 6.90E-27 | 1.35 [1.24-1.47] | 4.91E-12 |
| IBS-C | MACEs | 1.33 [1.17-1.51] | 1.33E-05 | 1.33 [1.17-1.51] | 1.64E-05 | 1.18 [1.03-1.35] | 1.39E-02 |
| | ACS | 1.49 [1.28-1.73] | 4.13E-07 | 1.48 [1.27-1.73] | 4.94E-07 | 1.32 [1.12-1.55] | 7.87E-04 |
| | Ischemic stroke | 1.02 [0.79-1.32] | 8.74E-01 | 1.02 [0.79-1.31] | 9.05E-01 | 0.92 [0.71-1.19] | 5.25E-01 |
| | HF | 1.17 [0.91-1.5] | 2.10E-01 | 1.17 [0.91-1.5] | 2.19E-01 | 1.06 [0.83-1.36] | 6.45E-01 |
| Functional constipation | MACEs | 1.1 [1.04-1.17] | 8.60E-04 | 1.1 [1.04-1.16] | 1.04E-03 | 1.09 [1.03-1.16] | 4.93E-03 |
| | ACS | 1.12 [1.05-1.2] | 1.28E-03 | 1.12 [1.04-1.2] | 1.42E-03 | 1.1 [1.02-1.18] | 1.03E-02 |
| | Ischemic stroke | 1.05 [0.95-1.17] | 3.37E-01 | 1.05 [0.94-1.17] | 3.62E-01 | 1.04 [0.94-1.16] | 4.54E-01 |
| | HF | 0.98 [0.88-1.09] | 7.08E-01 | 0.98 [0.88-1.09] | 6.87E-01 | 0.98 [0.88-1.09] | 7.01E-01 |

**Legend**: MACEs: Major adverse cardiac events; ACS: Acute coronary syndrome; HF: Heart failure; CCBs: Calcium channel blockers; BMI: Body mass index; CI: Confidence interval; SMuRFs: Standard modifiable cardiovascular risk factors, including hypertension, diabetes, smoking status, and hypercholesterolemia. In red, $P<0.05$.

**Supplementary Table 5.** Publicly available GWAS summary statistics for ACS, ischemic stroke, and HF.

| **Phenotype** | **N cases** | **N controls** | **$h^2_{SNP}$** | **N GWAS loci** | **Study** |
|---|---|---|---|---|---|
| Acute coronary syndrome (ACS) | 61,000 | 577,000 | 0.094 | 80 | Hartiala JA, et al.. Eur Heart J. 2021 Mar 1;42(9):919-933 |
| Ischemic stroke | 67,162 | 454,450 | 0.026 | 32 | Malik R, et al.. Nat Genet. 2018 Apr;50(4):524-537 |
| Heart failure (HF) | 47,309 | 930,014 | 0.035 | 11 | Shah S, et al.. Nat Commun. 2020 Jan 9;11(1):163. |

**Legend**: h2SNP: SNP-based heritability; GWAS: Genome-wide association study.
